# Haemochromatosis genetic variants and musculoskeletal outcomes: 11.5 year follow-up in the UK Biobank cohort study

**DOI:** 10.1101/2022.12.16.22283577

**Authors:** Lucy R Banfield, Karen M Knapp, Luke C Pilling, David Melzer, Janice L Atkins

## Abstract

**Background:** The iron overload disorder haemochromatosis is primarily caused by the homozygous *HFE* p.C282Y variant, but the scale of excess related musculoskeletal morbidity is uncertain.

**Methods:** We estimated haemochromatosis-genotype associations with clinically diagnosed musculoskeletal outcomes and joint replacement surgeries in the UK Biobank community cohort. 451,143 European ancestry participants (40-70 years at baseline) were followed in hospital records (mean 11.5 years). Cox proportional hazards models estimated *HFE* p.C282Y and p.H63D associations with incident outcomes.

**Results:** Male p.C282Y homozygotes (n=1,294) had increased incidence of osteoarthritis (n=52, HR: 2.12 [95% CI:1.61-2.80]; p=8.8*10^-8)^, hip replacement (n=88, HR:1.84 [95% CI: 1.49-2.27]; p=1.6*10^-8^), knee replacement (n=61, HR:1.54 [95% CI:1.20-1.98]; p=8.4*10^-4^), ankle and shoulder replacement, compared to males with no *HFE* mutations. Cumulative incidence analysis, using Kaplan-Meier lifetable probabilities demonstrated 10.4% of male homozygotes were projected to develop osteoarthritis and 15.5% to have hip replacements by age 75, versus 5.0% and 8.7% respectively without mutations. Male p.C282Y homozygotes also had increased incidence of femoral fractures (n=15, HR:1.72 [95% CI: 1.03-2.87]; p=0.04) and osteoporosis (n=21, HR:1.71 [95% CI: 1.11-2.64, p=0.02), although the latter association was limited to those with liver fibrosis/cirrhosis diagnoses. Female p.C282Y homozygotes had increased incidence of osteoarthritis only (n=57, HR:1.46, [95% CI: 1.12-1.89]; p=0.01). Male p.C282Y/p.H63D compound heterozygotes experienced a modest increased risk of hip replacements (n= 234, HR: 1.17 [95% CI: 1.02–1.33] p=0.02), but this did not pass multiple testing corrections.

**Conclusions:** In this large community cohort, the p.C282Y homozygote genotype was associated with substantial excess musculoskeletal morbidity in males. Wider *HFE* genotype testing may be justified, including in orthopaedic clinics serving higher *HFE* variant prevalence populations.

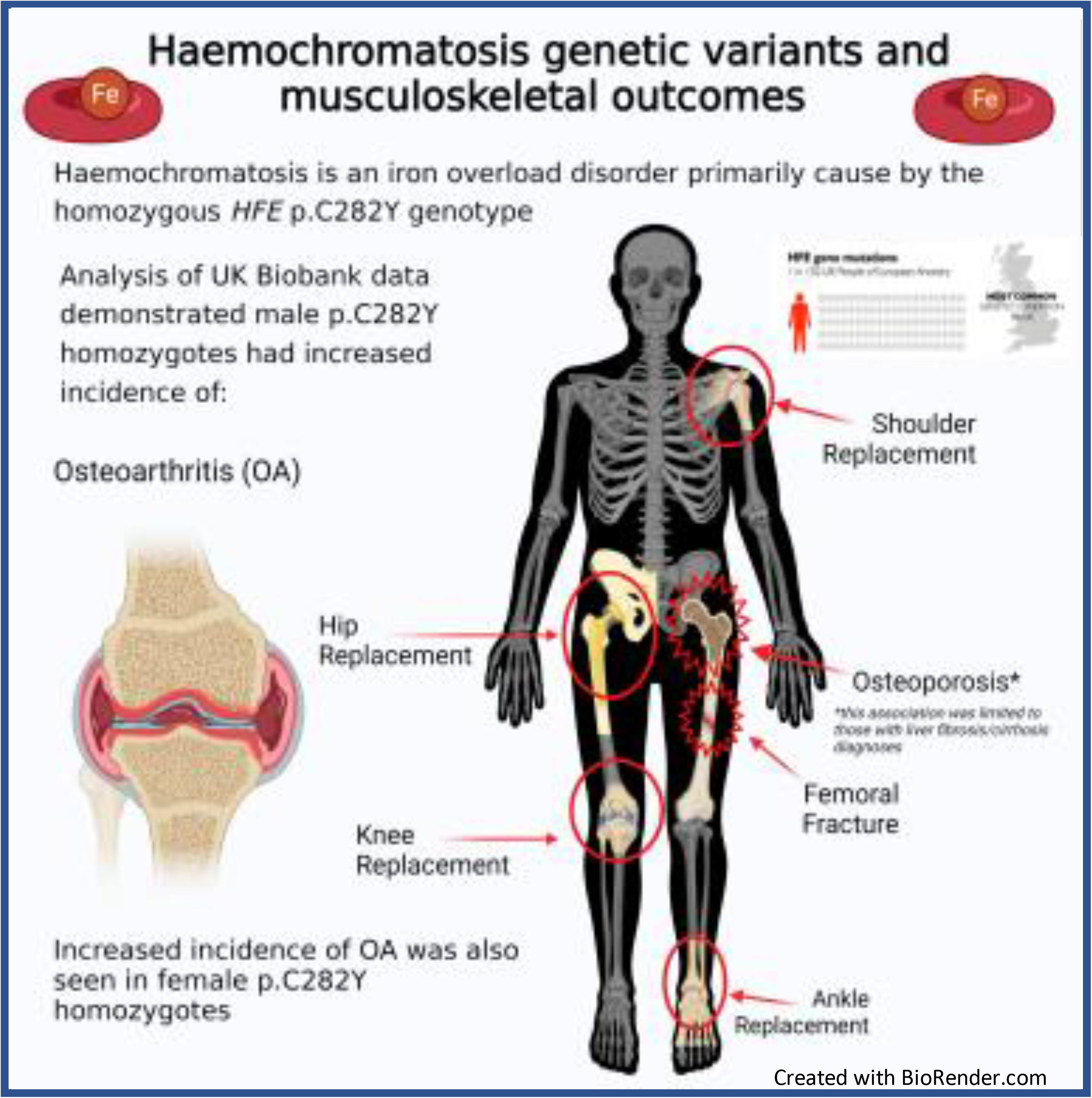

## Introduction

The iron overload disorder haemochromatosis is an autosomal recessive condition predominantly caused by mutations in the *HFE* gene, particularly the p.C282Y mutation, and to a lesser extent the p.H63D mutation. The p.C282Y mutation can cause accumulation of iron which builds up over time and is associated with osteoarthritis (OA), diabetes, liver disease, liver cancer as well as other conditions,^1–3^ although only a minority of community populations with the p.C282Y homozygote genotype actually develop associated clinical disease: i.e. the clinical penetrance is limited. The musculoskeletal impact of haemochromatosis has been widely reported and it has long been recognised that patients with haemochromatosis experience joint pain at a relatively young age.^4^ In addition, the progression to joint replacement surgery in this population has also been reported. Previous analysis of UK Biobank data^2^ (mean 7-year follow-up) found that in male p.C282Y homozygotes, the odds of hip replacement were raised (OR:2.62 [95% CI:1.97-3.49]; *P*<0.001). This is a similar incidence to a previous study which found a tripled risk of hip replacement in haemochromatosis patients.^5^

Another widely reported musculoskeletal impact of haemochromatosis is osteoporosis (OP), with evidence suggesting approximately 25% of patients with haemochromatosis will suffer from OP,^6^ and an even greater number from osteopaenia.^7^ A sequela of OP is the risk of fracture and researchers have observed higher risks within haemochromatosis patients. Early case studies presented unusual fractures in relatively young patients with markedly low bone mineral density (BMD) on dual-energy x-ray absorptiometry (DXA) scanning ranging from −3 to −5 SD’s in the lumbar spine ^8, 9^. While increased odds of wrist fracture have been demonstrated in haemochromatosis patients (OR:1.7, 95% CI:1.0-3.0, p=0.04),^10^ the association of wider fragility fracture presentation has not been proven.^11^

Although excess musculoskeletal disease and associated surgical procedures clearly occur in clinically diagnosed haemochromatosis patients, less is known about outcomes in community genotyped cohorts, especially to the older ages when joint replacements are common. Follow-up data over 11.5 years from the largest study of community dwelling participants who have been genotyped for *HFE* p.C282Y and p.H63D status, was used to examine associations between haemochromatosis genotypes and musculoskeletal outcomes. The current analysis of UK Biobank data advances our previous analyses ^2^ by including longer follow-up and a wider range of musculoskeletal outcomes.

## Methods

### UK Biobank participants

The UK Biobank study includes 502,464 volunteers aged 40 to 70 years. Baseline assessments (2006-10) included demographics, lifestyle, disease history, physiological measurements and collection of blood for genotyping^12^. Genotyping data were available for 451,143 participants of European ancestry with *HFE* p.C282Y (rs1800562) and *HFE* p.H63D (rs1799945) genotype information (see Supplementary Methods for details). Participants were followed-up via electronic hospital admissions data until 2020. UK Biobank participants were notified of relevant health related findings at the time of the baseline assessment. However, participants are not notified of subsequent findings, including genotyping.

### Baseline variables

Prevalent disease diagnoses were derived from self-reports at baseline though a verbal interview with a trained nurse, plus hospital inpatient data (National Health Service Hospital Episode Statistics) from 1996 to the baseline assessment, coded using International Classification of Diseases, 10th Revision (ICD-10). We examined a range of prevalent musculoskeletal outcomes, plus prevalent diagnoses of haemochromatosis or liver cirrhosis/fibrosis. Height and weight were measured at baseline, and body mass index (BMI) was calculated in kg/m^2^. Serum 25-hydroxyvitamin D (25[OH]D, a proxy for vitamin D) was measured in nanomoles per liter (nmol/L). Participants (n= 259,529) had bilateral calcaneal Quantitative Ultrasound Scanning (QUS) measurements obtained the Sahara Clinical Bone Sonometer (Hologic, Inc., Marlborough, MA, USA). Broadband ultrasound attenuation (BUA) and speed of sound (SOS) were derived, and combined to calculate the quantitative ultrasound index (QUI), any preventative maintenance, routine servicing and calibration of equipment within Biobank assessment centres is managed by the centre manager. ^13^

### Incident disease outcomes

Incident disease diagnoses, operations and procedures were identified during follow-up in hospital inpatient data, from baseline assessment to December 2020. Disease outcomes were coded using ICD-10. Operations and procedures were coded using the OPCS Classification of Interventions and Procedures version 4 (OPCS-4) (see Supplementary Table 1). Incident musculoskeletal outcomes examined included osteoarthritis (OA), hip/knee/shoulder or ankle replacement surgery, osteoporosis (OP), any reported fracture, femoral fracture (including neck of femur) and wrist fracture.

### Statistical analysis

Cox proportional hazards regression tested associations between genotypes and risk of incident musculoskeletal diagnoses/surgical procedures. Results are presented in each of the p.C282Y-H63D genotype groups, compared to those without mutations. Incident outcomes excluded participants with each baseline prevalent diagnosis/surgical procedure (from self-reports at baseline, plus hospital inpatient data from 1996 to baseline). Models were stratified by sex and adjusted for age, assessment centre, genotyping array, and 10 genetic principal components generated in participants of European descent, accounting for population genetics substructure. Excess musculoskeletal outcomes are well documented in p.C282Y homozygotes. However, for other genotype groups we performed multiple testing correction (Bonferroni-corrected p-value of 0.006, dividing the 0.05 significance level by 9 musculoskeletal outcomes). Cumulative incidence analysis, using Kaplan-Meier lifetable probabilities estimated hypothetical cumulative incident case numbers from age 40 to 75 years, in 5-year bands by genotype and sex. Proportional hazards assumptions for the musculoskeletal outcomes and replacement surgeries were tested with Schoenfeld residuals, which did not indicate violation of the assumption. Analyses were performed in Stata 15.1

### Sensitivity analysis

Previous literature questioned the impact of liver disease on bone quality in haemochromatosis patients^7, 14^ so a sensitivity analysis of the association between genotypes and OP and femoral fracture were performed excluding participants with a baseline liver fibrosis/cirrhosis diagnosis, to determine any relationship between liver disease and bone fragility. We also excluded participants with a vitamin D deficiency (<25nmol/L).^15^

To assess the impact of clinical haemochromatosis on musculoskeletal outcomes, a sensitivity analysis excluding those participants with diagnosed haemochromatosis at baseline was performed. In addition, we also excluded participants who had a hip fracture code recorded within 5 days prior to the replacement surgery to determine rates of hip replacement surgery unrelated to trauma. Since a prior fracture significantly increases the risk of a subsequent fracture, we carried out additional sensitivity analysis, using logistic regression to assess the odds of ever having a fracture by haemochromatosis genotype (combining baseline self-reported data and follow-up data from hospital records).

Primary care follow-up data were available in a subset of participants (n=209,795) from baseline until 2016-2017. Incident outcomes likely to be recorded in a primary care setting (OA and OP) were categorized from primary care ‘Read codes’ (see Supplementary Table 2).^16^ In this subset of participants with primary care data, Cox proportional hazards regression were repeated to test association between genotypes and the risk of OA and OP (combining diagnoses in hospital inpatient records or primary care data).

For incident outcomes, we also used Fine and Gray competing risk models, including death prior to diagnosis of the outcome as a competing risk event (follow-up data available via death records to December 2020). To check whether results are biased due to inclusion of related participants we repeated the primary analyses randomly excluding one of each pair of participants related to the third degree or closer identified by KING kinship analysis^17^ (381,000 participants remaining).

## Results

### Characteristics of participants

Analyses included 451,143 European descent participants aged 40 to 70 years at baseline, 54% of the sample were women and mean age was 56.8 years (SD 8.0). The mean follow-up period was 11.5 years (max 14.8). There were 2,890 p.C282Y homozygotes (0.6%), with 12.1% of male (156/1,294) and 3.4% of female (54/1,596) p.C282Y homozygotes diagnosed with haemochromatosis at baseline, increasing to 406 males (31.4%) and 302 females (18.9%) by the end of follow-up. Mean BMI was slightly lower in male p.C282Y homozygotes compared to those without mutations (27.6 vs 27.9 kg/m^2^, p=0.02), but did not differ in females (26.9 vs 27 kg/m^2^, p=0.21). Both male and female p.C282Y homozygotes had slightly lower mean vitamin D levels compared to those with no mutations (p=0.006 and p=0.01 respectively). There were no differences in BMD QUI scores in either male (p=0.07) or female (p=0.80) p.C282Y homozygotes compared to those without mutations. Male p.C282Y homozygotes had higher odds of baseline liver fibrosis/cirrhosis compared to those without mutations (1.7% vs 0.12%, OR:13.41, 95% CI:8.49-21.17, p=8.5*10^-29^), but this was not observed in females (0.25% vs 0.11% OR:2.09, 95% CI:0.77-5.65, p=0.15) (Table 1).

**Table 1.** Baseline characteristics of UK Biobank participants by p.C282Y/H63D genotypes and sex.

| **Male** |  |  |  |  |  |  |
| --- | --- | --- | --- | --- | --- | --- |
|  | **p.C282Y**<br>**Homozygotes** | **p.C282Y**<br>**Heterozygotes** | **Compound**<br>**Heterozygotes** | **H63D**<br>**Homozygotes** | **H63D**<br>**Heterozygotes** | **No mutations** |
| N (%) | 1,294 (0.6) | 24,575 (11.9) | 4,954 (2.4) | 4,674 (2.3) | 47,988 (23.3) | 122,845 (59.5) |
| Diagnosed haemochromatosis, N (%) | 156 (12.1) | 27 (0.1) | 29 (0.6) | 8 (0.2) | 17 (0.4) | 30 (0.2) |
| Mean age, years (SD) | 56.8 (8.2) | 57.0 (8.1) | 57.0 (8.1) | 57.0 (8.1) | 57.0 (8.1) | 57.0 (8.1) |
| Mean QUI (SD)* | 101.1 (23.5) | 101.9(23.6) | 102.3(22.3) | 101.9(23.3) | 102.5(23.3) | 102.6(23.7) |
| Mean BMI, kg/m<sup>2</sup> (SD) | 27.6 (4.1) | 27.9 (4.2) | 27.8 (4.2) | 27.9 (4.2) | 27.9 (4.3) | 27.9 (4.2) |
| Mean Vit D, nmol/L (SD) | 47.7 (19.6) | 49.5 (21.2) | 48.9 (21.2) | 49.2 (21) | 49.6 (21) | 49.8 (21.1) |
| Fibrosis/Cirrhosis (%) | 22 (1.7) | 36 (0.2) | 6 (0.1) | <5 (<0.1) | 75 (0.2) | 145 (0.1) |

| **Female** |  |  |  |  |  |  |
| --- | --- | --- | --- | --- | --- | --- |
|  | **p.C282y**<br>**Homozygotes** | **p.C282y**<br>**Heterozygotes** | **Compound**<br>**Heterozygotes** | **H63D**<br>**Homozygotes** | **H63D**<br>**Heterozygotes** | **No mutations** |
| N (%) | 1,596 (0.7) | 29,157 (11.9) | 5,745 (2.3) | 5,584 (2.3) | 57,023 (23.3) | 145,708 (59.5) |
| Diagnosed haemochromatosis, N (%) | 54 (3.4) | 5 (0.02) | 12 (0.2) | <5 (<0.09) | 6 (0.01) | 8 (0.01) |
| Mean age, years (SD) | 56.9 (8.0) | 56.5 (8.0) | 56.5 (7.9) | 56.6 (8.1) | 56.6 (7.9) | 56.6 (7.9) |
| Mean QUI (SD)* | 93.4(19.5) | 93.5(18.9) | 94.0(18.6) | 93.4(18.6) | 93.9(19.0) | 93.5(18.9) |
| Mean BMI, kg/m<sup>2</sup> (SD) | 26.9 (5.1) | 27 (5.1) | 27.1 (5.2) | 27.1 (5.3) | 27 (5.1) | 27 (5.2) |
| Mean Vitamin D, nmol/L (SD) | 48.1 (20.3) | 49.2 (21) | 48.9 (20.7) | 49.3 (20.6) | 49.4 (21) | 49.8 (20.9) |
| Fibrosis/Cirrhosis (%) | <5 (<0.3) | 34 (0.1) | 7 (0.1) | 9 (0.2) | 67 (0.1) | 159 (0.1) |

\*Quantitative Ultrasound Index (QUI) data were available for a subset of n=259,529 participants.
QUI is a combined measurement including BUA and SOS, which provides an analogue of BMD.
Numbers presented are mean (SD) for continuous variables and N (%) for categorical variables.
BMD = bone mineral density; SD = standard deviation

### Hazard ratios for incident musculoskeletal outcomes

During follow up, both male (n=52, HR: 2.12 [95% CI: 1.61-2.80] p=8.8*10^-8^) and female (n=57 HR: 1.46 [95% CI: 1.12-1.89] p=0.01) p.C282Y homozygotes had increased risks of OA compared to those with neither variant. Our analysis found increased risks of joint replacement surgery in male p.C282Y homozygotes at the knee (n=61, HR: 1.54 [95% CI:1.2-1.98] p=8.4*10^-4^), hip (n=88, HR: 1.84 [95% CI:1.49–2.27] p=1.6*10^-8^), shoulder (n<5, HR: 7.55 [95% CI:1.79-31.86] p=0.006), and ankle (n=13, HR 20.08 [95% CI:10.96-36.78] p=2.7*10^-22^) compared to those without mutations. However, increased risks of these four replacement surgeries were not seen in female p.C282Y homozygotes (Table 2).

**Table 2.** Associations between p.C282Y/H63D genotypes and risk of osteoarthritis and joint replacements, by sex*.

|  |  | Osteoarthritis |  | Knee Replacement |  | Hip Replacement |  | Shoulder Replacement |  | Ankle Replacement |  |
| --- | --- | --- | --- | --- | --- | --- | --- | --- | --- | --- | --- |
|  |  | HR<br>(95% CI) | P Value | HR<br>(95% CI) | p Value | HR<br>(95% CI) | p Value | HR<br>(95% CI) | p Value | HR<br>(95% CI) | p Value |
| No genetic mutations | Male | n=2,445<br>1.00 |  | n=3,821<br>1.00 |  | n=5,013<br>1.001 |  | n=28<br>1.00 |  | n=60<br>1.00 |  |
|  | Female | n=3,568<br>1.00 |  | n=4,925<br>1.00 |  | n=7,946<br>1.00 |  | n=95<br>1.00 |  | n= 55<br>1.00 |  |
| p.C282y<br>Homozygotes | Male | n= 52<br>2.12<br>(1.61-2.80) | 8.8*10<sup>-8</sup> | n=61<br>1.54<br>(1.20-1.98) | 8.4*10<sup>-4</sup> | n=88<br>1.84<br>(1.49-2.27) | 1.6*10<sup>-8</sup> | n<5<br>7.55<br>(1.79-31.86) | 0.006 | n=13<br>20.08<br>(10.96-36.78) | 2.7*10<sup>-22</sup> |
|  | Female | n= 57<br>1.46<br>(1.12-1.89) | 0.01 | n=63<br>1.14<br>(0.89-1.46) | 0.32 | n=100<br>1.17<br>(0.96-1.42) | 0.12 | n<5<br>1.97<br>(0.49-8.04) | 0.34 | n<5<br>1.58<br>(0.22-11.42) | 0.65 |
| p.C282y<br>Heterozygotes | Male | n=511<br>1.02<br>(0.92-1.12) | 0.72 | n=841<br>1.08<br>(1.01-1.17) | 0.04 | n=1,083<br>1.09<br>(1.02-1.16) | 0.02 | n=5<br>0.92<br>(0.35-2.37) | 0.86 | n=16<br>1.30<br>(0.75-2.26) | 0.35 |
|  | Female | n= 753<br>1.05<br>(0.97-1.13) | 0.24 | n=1,001<br>1.02<br>(0.95-1.09) | 0.55 | n=1,535<br>0.98<br>(0.93-1.04) | 0.49 | n=15<br>0.82<br>(0.48-142) | 0.49 | n=6<br>0.53<br>(0.23-1.24) | 0.14 |
| Compound<br>Heterozygotes | Male | n=96<br>0.96<br>(0.78-1.17) | 0.67 | n=145<br>0.92<br>(0.78-1.09) | 0.34 | n=234<br>1.17<br>(1.02-1.33) | 0.02 | n=0<br>N/A | N/A | n<5<br>0.41<br>(0.06-2.96) | 0.38 |
|  | Female | n= 164<br>1.16<br>(0.99-1.36) | 0.06 | n=201<br>1.04<br>(0.91-1.20) | 0.54 | n=312<br>1.02<br>(0.91-1.14) | 0.72 | n<5<br>0.28<br>(0.04-2.03) | 0.21 | n<5<br>1.78<br>(0.65-4.93) | 0.26 |
| H63D<br>Homozygotes | Male | n=81<br>0.87<br>(0.70-1.08) | 0.21 | n=134<br>0.92<br>(0.77-1.09) | 0.33 | n=192<br>1.01<br>(0.88-1.17) | 0.85 | n<5<br>0.95<br>(0.13-6.95) | 0.96 | n<5<br>0.88<br>(0.21-3.60) | 0.86 |
|  | Female | n= 146<br>1.08<br>(0.91-1.27) | 0.88 | n=187<br>0.99<br>(0.86-1.15) | 0.90 | n=286<br>0.94<br>(0.83-1.06) | 0.29 | n<5<br>1.12<br>(0.41-3.04) | 0.83 | N<5<br>0.94<br>(0.23-3.87) | 0.94 |
| H63D<br>Heterozygotes | Male | n=989<br>1.03<br>(0.96-1.11) | 0.41 | n=1,548<br>1.03<br>(0.97-1.10) | 0.27 | n=1,945<br>0.99<br>(0.94-1.05) | 0.79 | n=13<br>1.19<br>(0.62-2.30) | 0.60 | n=37<br>1.59<br>(1.05-2.39) | 0.03 |
|  | Female | n=1,405<br>1.01<br>(0.95-1.07) | 0.32 | n=1,902<br>0.99<br>(0.94-1.04) | 0.71 | n=3,071<br>0.99<br>(0.95-1.04) | 0.74 | n=24<br>0.65<br>(0.42-1.02) | 0.06 | n=21<br>0.97<br>(0.59-1.61) | 0.91 |

\* HR (Hazard ratio) compared to those with neither HFE mutation. Cox proportional hazards regression models adjusted for age, assessment centre, genotyping array, and genetic principal components 1–10. Excluding prevalent disease diagnoses at baseline.

There was a modest increased risk of hip and knee replacement within male p.C282Y heterozygotes (knee: n=841, HR: 1.08 [95% CI: 1.01-1.17] p=0.04, hip: n=1,083 HR: 1.09 [95% CI: 1.02–1.16] p=0.02). Male p.C282Y/p.H63D compound heterozygotes also demonstrated a modest increased risk of hip replacement (n= 234, HR: 1.17 [95% CI: 1.02–1.33] p=0.02) and male p.H63D heterozygotes had a modest increased risk of ankle replacement (n=37, HR: 1.59 [95% CI: 1.05-2.39) p=0.03). However, following Bonferroni correction for multiple testing, none of the above findings in p.C282Y heterozygous, p.H63D heterozygous or p.C282Y/p.H63D compound heterozygotes remained significant.

Analysis also demonstrated an increased risk of OP in male p.C282Y homozygotes (n=21, HR: 1.71 [95% CI: 1.11-2.64] p=0.02) and femoral fractures (n=15, HR: 1.72 [95% CI: 1.03-2.87] p=0.04). However, male p.C282Y homozygotes did not demonstrate increased risks of ‘any reported fractures or wrist fractures, although number of wrist fractures was low at n=<5. The incidence of OP and fractures in female p.C282Y homozygotes when compared to those with no mutations was not increased (Table 3).

**Table 3.** Associations between p.C282Y/H63D genotypes and risk of osteoporosis and fractures, by sex*.

|  |  | Osteoporosis |  | Femoral Fracture<br>(inc. neck of femur) |  | Wrist Fracture |  | Any Reported fracture |  |
| --- | --- | --- | --- | --- | --- | --- | --- | --- | --- |
|  |  | HR<br>(95% CI) | P Value | HR<br>(95% CI) | P Value | HR<br>(95% CI) | P Value | HR<br>(95% CI) | P Value |
| No genetic mutations | Male | n=1,181<br>1.00 |  | n=824<br>1.00 |  | n= 477<br>1.00 |  | n=5,085<br>1.00 |  |
|  | Female | n=5,357<br>1.00 |  | n=1,663<br>1.00 |  | n= 2,640<br>1.00 |  | n=8,886<br>1.00 |  |
| p.C282y Homozygotes | Male | n=21<br>1.71<br>(1.11-2.64) | 0.02 | n= 15<br>1.72<br>(1.03-2.87) | 0.04 | n<5<br>0.59<br>(0.19-1.83) | 0.36 | n= 66<br>1.24<br>(0.97-1.58) | 0.09 |
|  | Female | n=71<br>1.16<br>(0.92-1.47) | 0.21 | n= 18<br>0.92<br>(0.58-1.46) | 0.72 | n= 32<br>1.05<br>(0.74-1.48) | 0.79 | n= 105<br>1.02<br>(0.85-1.24) | 0.80 |
| p.C282y Heterozygotes | Male | n=242<br>1.01<br>(0.88-1.60) | 0.89 | n= 179<br>1.07<br>(0.426) | 0.43 | n= 82<br>0.85<br>(0.68 – 1.08) | 0.19 | n= 1,013<br>0.98<br>(0.92-1.05) | 0.62 |
|  | Female | n=1,034<br>0.97<br>(0.90-1.03) | 0.30 | n= 335<br>1.01<br>(0.90-1.13) | 0.91 | n= 552<br>1.03<br>(0.94-1.13) | 0.52 | n= 1,814<br>1.01<br>(0.92-1.06) | 0.64 |
| Compound Heterozygotes | Male | n=51<br>1.05<br>(0.80-1.4) | 0.69 | n= 39<br>1.15<br>(0.83-1.59) | 0.39 | n= 19<br>0.98<br>(0.62-1.55) | 0.93 | n= 197<br>0.95<br>(0.82-1.10) | 0.49 |
|  | Female | n=188<br>0.90<br>(0.78-1.04) | 0.14 | n= 72<br>1.11<br>(0.88-1.40) | 0.39 | n= 109<br>1.03<br>(0.85-1.25) | 0.76 | n= 350<br>0.99<br>(0.89-1.10) | 0.82 |
| H63D Homozygotes | Male | n=38<br>0.84<br>(.61-1.17) | 0.32 | n= 32<br>1.02<br>(0.72-1.45) | 0.91 | n= 8<br>0.44<br>(0.22-0.89) | 0.02 | n= 189<br>0.98<br>(0.84-1.13) | 0.75 |
|  | Female | n=200<br>0.96<br>(0.84-1.11) | 0.61 | n= 54<br>0.83<br>(0.64-1.09) | 0.19 | n= 88<br>0.86<br>(0.70-1.07) | 0.18 | n= 311<br>0.91<br>(0.81-1.02) | 0.10 |
| H63D Heterozygotes | Male | n=457<br>0.99<br>(0.89-1.10) | 0.84 | n= 330<br>1.02<br>(0.90-1.16) | 0.74 | n= 167<br>0.90<br>(0.75-1.07) | 0.23 | n= 1,942<br>0.98<br>(0.93-1.03) | 0.38 |
|  | Female | n=2,017<br>0.96<br>(0.92-1.01) | 0.16 | n= 658<br>1.02<br>(0.93-1.11) | 0.71 | n= 1,058<br>1.02<br>(0.95-1.10) | 0.53 | n= 3,529<br>1.02<br>(0.98-1.06) | 0.45 |

HR (Hazard ratio) compared to those with neither HFE mutation. Cox proportional hazards regression models adjusted for age, assessment centre, genotyping array, and genetic principal components 1–10. Excluding prevalent operations/procedures at baseline.

### Cumulative incidence analysis of incident musculoskeletal outcomes during follow-up

In cumulative incidence analysis, using Kaplan-Meier lifetable probability estimates, 10.4% (95% CI: 7.8-13. 7) of male p.C282Y homozygotes were projected to develop OA by age 75, compared to 5.0% (95% CI: 4.7-5.3) in those without mutations (excess proportion =5.4%). For hip replacement, 15.5% (95% CI: 12.6-19.1) of male p.C282Y homozygotes were projected to undergo hip replacement surgery, compared to 8.7% (95% CI: 8.5-9.0) of those without mutations (excess proportion =6.8%). In male p.C282Y homozygotes, by age 75 9.6% (95% CI: 7.4-12.4) were projected to undergo knee replacement surgery compared to 6.6% (95% CI: 6.3-6.8) in those without mutations (excess proportion of 3.0%).

Overall, the numbers for both shoulder and ankle replacement surgeries were low and only small excess proportions were identified; by age 75 male p.C282Y homozygotes were projected to have an excess of 0.4% of shoulder replacement and 1.8% excess of ankle replacements compared to those with no genetic variation. When comparing genotypes relative to risk of incidence rates for OP, the differences were relatively small; projected excess of 1.5% by age 75 and 1% excess in male P.C282Y homozygotes (see Tables 4, 5 and 6).

**Table 4.**
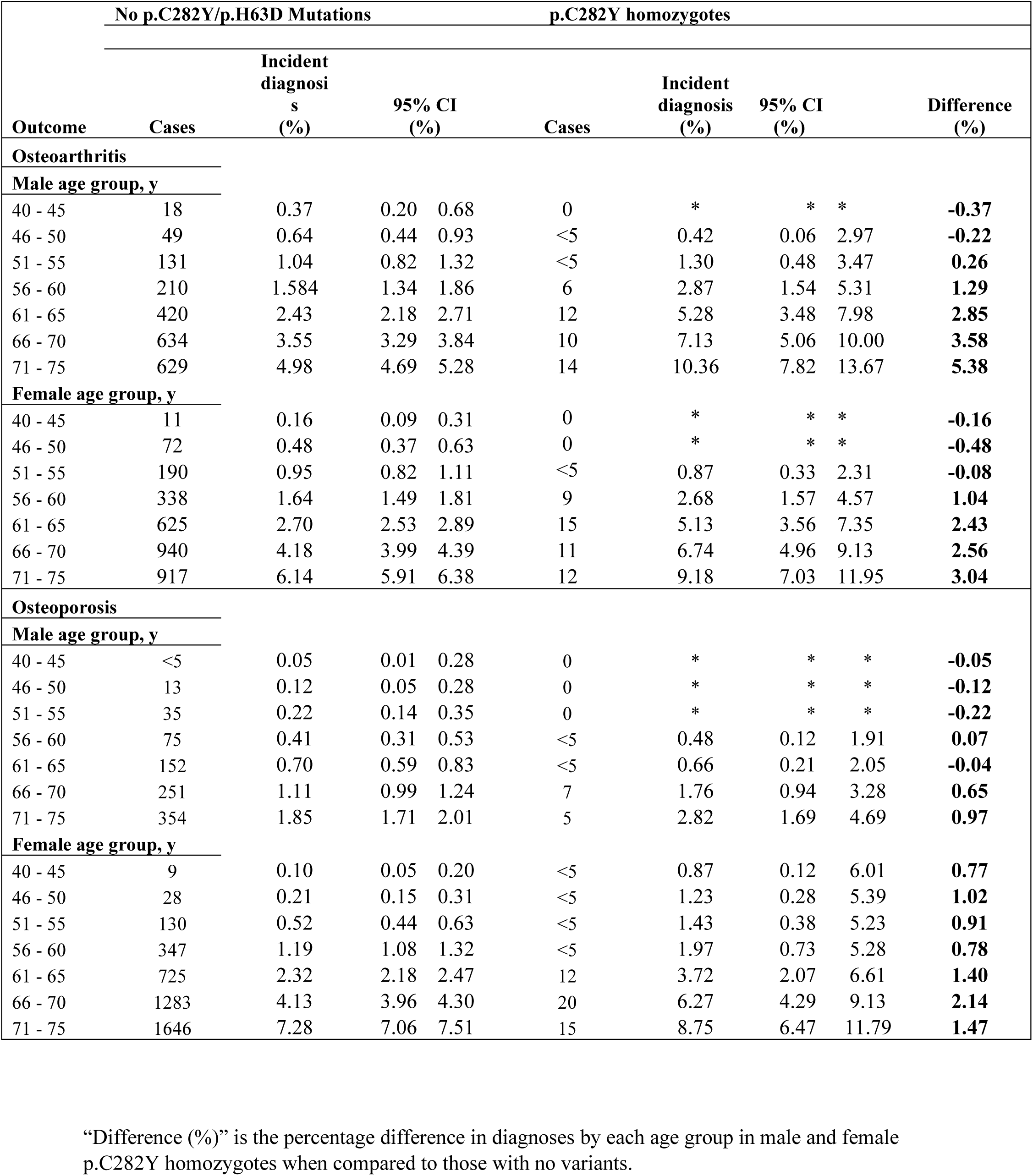
Cumulative incidence analysis, using Kaplan-Meier lifetable probability estimates of musculoskeletal outcomes by sex and *HFE* genotypes (p.C282Y homozygotes vs no mutations)

**Table 5.**
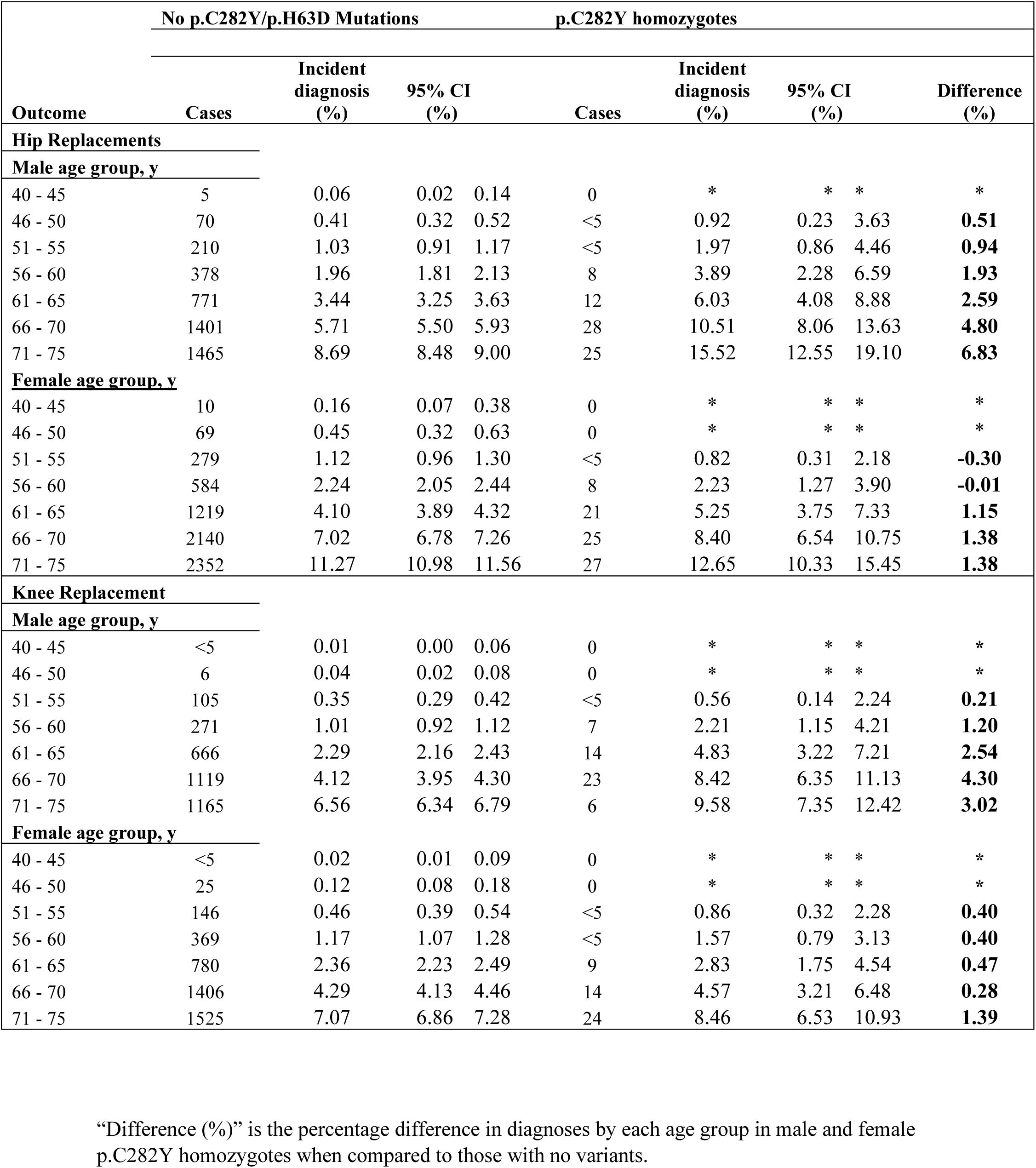
Cumulative incidence analysis, using Kaplan-Meier lifetable probability estimates of hip and knee joint replacement surgery by sex and *HFE* genotypes (p.C282Y homozygotes vs no mutations)

| No p.C282Y/p.H63D Mutations |  |  |  |  | p.C282Y homozygotes |  |  |  |  |
| --- | --- | --- | --- | --- | --- | --- | --- | --- | --- |
| Outcome | Cases | Incident diagnosis (%) | 95% CI (%) |  | Cases | Incident diagnosis (%) | 95% CI (%) |  | Difference (%) |
| Hip Replacements |  |  |  |  |  |  |  |  |  |
| Male age group, y |  |  |  |  |  |  |  |  |  |
| 40 - 45 | 5 | 0.06 | 0.02 | 0.14 | 0 | * | * | * | * |
| 46 - 50 | 70 | 0.41 | 0.32 | 0.52 | <5 | 0.92 | 0.23 | 3.63 | 0.51 |
| 51 - 55 | 210 | 1.03 | 0.91 | 1.17 | <5 | 1.97 | 0.86 | 4.46 | 0.94 |
| 56 - 60 | 378 | 1.96 | 1.81 | 2.13 | 8 | 3.89 | 2.28 | 6.59 | 1.93 |
| 61 - 65 | 771 | 3.44 | 3.25 | 3.63 | 12 | 6.03 | 4.08 | 8.88 | 2.59 |
| 66 - 70 | 1401 | 5.71 | 5.50 | 5.93 | 28 | 10.51 | 8.06 | 13.63 | 4.80 |
| 71 - 75 | 1465 | 8.69 | 8.48 | 9.00 | 25 | 15.52 | 12.55 | 19.10 | 6.83 |
| Female age group, y |  |  |  |  |  |  |  |  |  |
| 40 - 45 | 10 | 0.16 | 0.07 | 0.38 | 0 | * | * | * | * |
| 46 - 50 | 69 | 0.45 | 0.32 | 0.63 | 0 | * | * | * | * |
| 51 - 55 | 279 | 1.12 | 0.96 | 1.30 | <5 | 0.82 | 0.31 | 2.18 | -0.30 |
| 56 - 60 | 584 | 2.24 | 2.05 | 2.44 | 8 | 2.23 | 1.27 | 3.90 | -0.01 |
| 61 - 65 | 1219 | 4.10 | 3.89 | 4.32 | 21 | 5.25 | 3.75 | 7.33 | 1.15 |
| 66 - 70 | 2140 | 7.02 | 6.78 | 7.26 | 25 | 8.40 | 6.54 | 10.75 | 1.38 |
| 71 - 75 | 2352 | 11.27 | 10.98 | 11.56 | 27 | 12.65 | 10.33 | 15.45 | 1.38 |
| Knee Replacement |  |  |  |  |  |  |  |  |  |
| Male age group, y |  |  |  |  |  |  |  |  |  |
| 40 - 45 | <5 | 0.01 | 0.00 | 0.06 | 0 | * | * | * | * |
| 46 - 50 | 6 | 0.04 | 0.02 | 0.08 | 0 | * | * | * | * |
| 51 - 55 | 105 | 0.35 | 0.29 | 0.42 | <5 | 0.56 | 0.14 | 2.24 | 0.21 |
| 56 - 60 | 271 | 1.01 | 0.92 | 1.12 | 7 | 2.21 | 1.15 | 4.21 | 1.20 |
| 61 - 65 | 666 | 2.29 | 2.16 | 2.43 | 14 | 4.83 | 3.22 | 7.21 | 2.54 |
| 66 - 70 | 1119 | 4.12 | 3.95 | 4.30 | 23 | 8.42 | 6.35 | 11.13 | 4.30 |
| 71 - 75 | 1165 | 6.56 | 6.34 | 6.79 | 6 | 9.58 | 7.35 | 12.42 | 3.02 |
| Female age group, y |  |  |  |  |  |  |  |  |  |
| 40 - 45 | <5 | 0.02 | 0.01 | 0.09 | 0 | * | * | * | * |
| 46 - 50 | 25 | 0.12 | 0.08 | 0.18 | 0 | * | * | * | * |
| 51 - 55 | 146 | 0.46 | 0.39 | 0.54 | <5 | 0.86 | 0.32 | 2.28 | 0.40 |
| 56 - 60 | 369 | 1.17 | 1.07 | 1.28 | <5 | 1.57 | 0.79 | 3.13 | 0.40 |
| 61 - 65 | 780 | 2.36 | 2.23 | 2.49 | 9 | 2.83 | 1.75 | 4.54 | 0.47 |
| 66 - 70 | 1406 | 4.29 | 4.13 | 4.46 | 14 | 4.57 | 3.21 | 6.48 | 0.28 |
| 71 - 75 | 1525 | 7.07 | 6.86 | 7.28 | 24 | 8.46 | 6.53 | 10.93 | 1.39 |

“Difference (%)” is the percentage difference in diagnoses by each age group in male and female p.C282Y homozygotes when compared to those with no variants.

**Table 6.** Cumulative incidence analysis, using Kaplan-Meier lifetable probability estimates of shoulder and ankle joint replacement surgery by sex and *HFE* genotypes (p.C282Y homozygotes vs no mutations)

| No p.C282Y/p.H63D Mutations |  |  |  |  | p.C282Y homozygotes |  |  |  |  |
| --- | --- | --- | --- | --- | --- | --- | --- | --- | --- |
| Outcome | Cases | Incident diagnosis (%) | 95% CI (%) |  | Cases | Incident diagnosis (%) | 95% CI (%) |  | Difference (%) |
| Shoulder replacements |  |  |  |  |  |  |  |  |  |
| Male age group, y |  |  |  |  |  |  |  |  |  |
| 40 - 45 | 0 | * | * | * | 0 | * | * | * | * |
| 46 - 50 | <5 | 0.01 | 0.00 | 0.04 | 0 | * | * | * | * |
| 51 - 55 | 0 | 0.01 | 0.00 | 0.04 | 0 | * | * | * | * |
| 56 - 60 | <5 | 0.01 | 0.00 | 0.04 | <5 | 0.25 | 0.04 | 1.77 | 0.24 |
| 61 - 65 | 5 | 0.02 | 0.01 | 0.05 | <5 | 0.42 | 0.10 | 1.72 | 0.40 |
| 66 - 70 | <5 | 0.03 | 0.02 | 0.05 | 0 | 0.42 | 0.10 | 1.72 | 0.39 |
| 71 - 75 | 9 | 0.05 | 0.03 | 0.07 | 0 | 0.42 | 0.10 | 1.72 | 0.37 |
| Female age group, y |  |  |  |  |  |  |  |  |  |
| 40 - 45 | 0 | * | * | * | 0 | * | * | * | * |
| 46 - 50 | <5 | * | 0.00 | 0.02 | 0 | * | * | * | * |
| 51 - 55 | <5 | 0.01 | 0.00 | 0.02 | 0 | * | * | * | * |
| 56 - 60 | <5 | 0.01 | 0.00 | 0.03 | 0 | * | * | * | * |
| 61 - 65 | 12 | 0.03 | 0.02 | 0.05 | 0 | * | * | * | * |
| 66 - 70 | 18 | 0.06 | 0.04 | 0.08 | <5 | 0.12 | 0.02 | 0.88 | 0.06 |
| 71 - 75 | 33 | 0.12 | 0.09 | 0.15 | <5 | 0.28 | 0.07 | 1.14 | 0.16 |
| Ankle replacements |  |  |  |  |  |  |  |  |  |
| Male age group, y |  |  |  |  |  |  |  |  |  |
| 40 - 45 | 0 | * | * | * | 0 | * | * | * | * |
| 46 - 50 | 0 | * | * | * | 0 | * | * | * | * |
| 51 - 55 | <5 | 0.00 | 0.00 | 0.02 | 0 | * | * | * | * |
| 56 - 60 | <5 | 0.01 | 0.00 | 0.03 | 0 | * | * | * | * |
| 61 - 65 | <5 | 0.02 | 0.01 | 0.04 | <5 | 0.58 | 0.19 | 1.78 | 0.56 |
| 66 - 70 | 24 | 0.06 | 0.04 | 0.08 | 5 | 1.36 | 0.68 | 2.71 | 1.30 |
| 71 - 75 | 16 | 0.09 | 0.07 | 0.13 | <5 | 1.90 | 0.15 | 3.41 | 1.81 |
| Female age group, y |  |  |  |  |  |  |  |  |  |
| 40 - 45 | 0 | * | * | * | 0 | * | * | * | * |
| 46 - 50 | 0 | * | * | * | 0 | * | * | * | * |
| 51 - 55 | <5 | 0.01 | 0.00 | 0.03 | 0 | * | * | * | * |
| 56 - 60 | 6 | 0.02 | 0.01 | 0.04 | 0 | * | * | * | * |
| 61 - 65 | 7 | 0.03 | 0.02 | 0.05 | 0 | * | * | * | * |
| 66 - 70 | 11 | 0.05 | 0.03 | 0.07 | <5 | 0.12 | 0.02 | 0.88 | 0.07 |
| 71 - 75 | 17 | 0.08 | 0.06 | 0.11 | 0 | 0.12 | 0.07 | 0.88 | 0.04 |

“Difference (%)” is the percentage difference in diagnoses by each age group in male and female p.C282Y homozygotes when compared to those with no variants.

### Sensitivity analyses

After excluding participants with a baseline diagnosis of fibrosis/cirrhosis (n=22), the association between male p.C282Y homozygosity and OP was no longer significant (HR: 1.41 [95% CI:0.87-2.28] p=0.16) appearing to indicate the liver fibrosis does impact on bone health. Similarly, after exclusion of those participants with a baseline diagnosis of haemochromatosis, the association between male p.C282Y homozygosity and OP was again weakened (HR; 1.20 [95% CI: 0.70 – 2.08] p=0.51) appearing to indicate that clinically evident haemochromatosis is linked with OP in male patients in particular. The associations between male p.C282Y homozygosity and OA, joint replacements and femoral fractures remained significant after the exclusion of those with a baseline haemochromatosis diagnosis. The increased risk of OP in male p.C282Y homozygotes remained when those with a vitamin D level of <25nmol/L (n=329 p.C282Y homozygotes) were excluded from analysis. The increased risk of hip replacement in male p.C282Y homozygotes remained after excluding hip fracture codes recorded within 5 days prior to the replacement surgery (n=16, HR: 1.84 [95% CI:1.48 – 2.2] p=3.9*10^-8^), appearing to indicate that OA is the most common causative factor for the hip arthroplasty seen within these male homozygotes.

In additional sensitivity analysis, examining the odds of ever having a fracture (baseline self-reports and hospital follow-up combined), male p.C282Y homozygotes had significantly increased odds of femoral fracture (OR: 2.03, 95% CI: 1.36-3.04, p = 0.001) and any reported fracture (OR: 1.32, 95% CI: 1.11-1.56, p = 0.001).

In the subset of participants with primary care data (n= 209,795), the association between male p.C282Y homozygosity and increased risk of OA (n=82, HR: 1.41 [95% CI:1.09-1.82] p=0.01) and OP (n=29, HR: 1.75 [95% CI:1.05-2.92] p=0.03) remained. However, the association between female p.C282Y homozygosity and increased risk of OA was no longer significant (n=105, HR:1.04 [95% CI:0.83-1.32] p=0.72).

In competing risk models, with death prior to the outcome as a competing risk, the association between male p.C282Y homozygosity and OA, joint replacements and OP, and the association between female p.C282Y homozygosity and OA, remained significant. However, the association between male p.C282Y homozygosity and femoral fractures was no longer significant (HR: 1.63 [0.99-2.67] p=0.06). We repeated primary analysis of OA and OP after excluding one of each pair of participants related to the third degree or closer, to check whether results are biased as related participants share genetic and environmental factors, but also in the context of haemochromatosis are more likely to be referred by screening independent of clinical severity. Unrelated male p.C282Y homozygotes still had increased likelihood of diagnoses of OA and OP compared to unrelated participants with no *HFE* p.C282Y or p.H63D alleles.

## Discussion

We used the UK Biobank cohort study (the largest community sample of haemochromatosis-genotype individuals to date) to estimate haemochromatosis-genotype associations with a range of musculoskeletal outcomes and joint replacement surgeries in 451,143 European ancestry participants. Over an 11.5-year follow-up period male p.C282Y homozygotes had increased risks of OA and joint replacement surgeries (hip, knee, shoulder and ankle), and increased risks of OP and femoral fracture (including neck of femur fracture) compared to those without mutations. These increased risks of OA, joint replacement and femoral fracture remained after excluding participants with diagnosed haemochromatosis at baseline. There was a modest increase in risk of hip and knee replacement within male p.C282Y heterozygotes, and hip replacement within male p.C282Y/p.H63D compound heterozygotes. However, there were no consistent increases in risk in osteoarthritis in these genotype groups and results were no longer significant after correction for multiple testing. Therefore, more work is needed to replicate these findings. The female p.C282Y homozygotes in our analysis only demonstrated an increased risk of OA compared to those with no genetic mutation. Our results provide the first estimates of cumulative risk to age 75 years; 10.4% of male p.C282Y homozygotes were projected to develop osteoarthritis and 15.5% to have hip replacements by age 75, versus 5.0% and 8.7% respectively without mutations.

The incidence of haemochromatosis related arthropathy is well established, with continued debate as to whether severity of iron overload is the main contributory factor, given the lack of evidence of any improvement in arthropathy symptoms, or slowing of disease progression following normalisation of iron levels.^18–20^ We recently demonstrated that male p.C282Y homozygotes carrying other alleles that increase serum iron levels had modestly increased likelihood of OA diagnosis,^21^ yet the effect is small and so the role of iron overload in OA onset remains to be fully elucidated. The male p.C282Y homozygotes in this community study demonstrated a twofold increased risk of developing OA, interestingly OA was the only musculoskeletal outcome analysed for this study that showed an increased risk in female p.C282Y homozygotes. Given that previous studies have demonstrated a potential under-reporting of OA diagnoses in primary care,^22^ this relationship may be an underestimate. In a large multicohort genome-wide association study (GWAS) meta-analysis of osteoarthritis, p.C282Y was a risk locus for hip OA, and effects were strongest in the males, despite the GWAS assuming an additive genotypic effect.^23^

The increased likelihood of patients with haemochromatosis requiring joint arthroplasty is another key area of research. It is thought that the excess iron in circulation may have a toxic effect on chondrocytes^24^ and this progressive degeneration results in an increased need for joint arthroplasty in order to relieve symptoms and improve quality of life. The increased likelihood of this surgery has been demonstrated in previous studies ^25^ (HR 2.88 [95% CI:2.39–3.43]) relative to primary hip arthroplasty, increasing when secondary hip replacements were assessed,^5^ echoing previous studies demonstrating an increased risk of homozygotes requiring bilateral total hip replacements (THR’s)^26^ in comparison to those with no p.C282Y mutation (OR: 5.86, 95% CI:2.36 – 14.57).^27^

Our analysis returned a hazard ratio of 1.84 (95% CI:1.49-2.27) for risk of hip replacement, reinforcing the evidence of greater risk for this type of surgery when compared to those without the genetic mutation. It did not show increased risk for female p.C282Y homozygotes in contradiction to an earlier study that identified a greater risk to young female homozygotes.^28^ In an apparent contradiction to our findings, a study by Oppl et al^29^ was unable to establish a relationship between *HFE* genotype, OA and hip joint replacement surgery. However, the mean age of their cohort was 59.4 years, and our cumulative incidence analysis data did not show a larger excess burden relative to OA until 66 years and older, with a similar picture when related to hip replacements, potentially explaining the differing conclusions.

Increased risks were seen with respect to knee arthroplasty in male p.C282Y homozygotes, again in line with previously published studies.^10, 28^ and also an increase in risk of ankle replacement surgery. This ankle surgery risk estimate is much larger than that reported in a previous meta-analysis (RR: 8.94, [95% CI:3.85-20.78]), although that was based on haemochromatosis diagnosis, not by genotype as in our current study.^30^ It may be worth noting that ankle replacement surgery is still a relatively specialist operation performed in highly specialized centres, ankle arthrodesis is a more likely procedure outside such centres, so our figures may underestimate true rate of surgical intervention.

Of additional relevance were increased risks relative to shoulder arthroplasty, again seen within male p.C282Y homozygotes. These increased risks have been reported in the literature, but primarily as case reports rather than larger scale cohort studies. A recent study determined that there was a trend towards shoulder arthroplasty ^31^ and although the number of shoulder replacement surgeries in the current study was small (n=2), this appears to be the first to identify an increased risk of shoulder arthroplasty associated with haemochromatosis-genotypes.

Evidence linking OP with haemochromatosis has been accumulating over a number of years, however the cause of this relationship is still a subject of debate. Experimental studies have suggested that iron overload inhibits osteoblastic activity and hydroxyapatite growth leading to reduced bone formation. Haemochromatosis is linked with liver disease, diabetes and hypogonadism, and although the latter is a rare complication, these conditions are all associated with bone loss,^10^ making a clear mechanism difficult to definitively establish. The two main factors that appear to impact on BMD relative to haemochromatosis are either severity of iron overload and/or liver cirrhosis, with BMI, alkaline phosphatase (ALP) levels and genetic status also potential contributory factors, however the published data is contradictory as to which of these is most strongly associated.^7, 32^ While studies have demonstrated increased risk of OP independent of cirrhosis and hypogonadism but still related to severity of iron overload, in contrast, another more recent cross-sectional analysis found that whilst bone fragility was observed in a fifth of patients with haemochromatosis, this was independent of the severity of iron overload and instead strongly associated with hepatic cirrhosis (OR 8.20 [95% CI 1.74-38.68], *P*= 0.008).^32^ In this cohort of male p.C282Y homozygotes there was an increased risk of incident OP and incident femoral fracture, and increased odds of ever having a femoral fracture and ever having any fracture.

The other contributory factor suggested as a potential cause of the OP seen in this patient group is liver disease. In our cohort of male p.C282Y homozygotes, when we excluded those with a baseline diagnosis of liver fibrosis and cirrhosis the association of the genotype with OP was no longer significant. This would appear to indicate that hepatocellular disease does contribute towards OP in the male p.C282Y homozygotes. Being diagnosed with haemochromatosis could lead to more investigations for osteoporosis, but this seems to be only a possibility within those with liver disease, given the attenuation of the estimate when the liver disease group is removed. After exclusion of participants with a baseline haemochromatosis diagnosis, the association between male p.C282Y homozygosity and OP was also no longer significant, suggesting that severity of iron overload may affect risk of OP.

Given the increased risk of osteoporosis, we also reviewed the BMD data by sex and by genotype to ascertain whether there were any notable differences between homozygotes and controls. There was no marked difference between homozygotes and other genotypes with the mean QUI remaining relatively comparable across all groups, albeit with the homozygotes appearing at the lower end of the range. These BMD results on the face of it appear to conflict with the increased risk of osteoporosis in the homozygotes and needs further investigation.

Although there were very slightly increased HR for females with respect to OP and fracture (any), these were not significant. With regard fracture, this analysis again demonstrated an increased risk for the male p.C282Y homozygotes in respect of femoral fracture, including fracture of the femoral neck, and increased odds of ever having a fracture. There have been several studies which indicate a link between haemochromatosis and fragility fracture,^8–10, 32, 33^ however this still remains an under-researched aspect of haemochromatosis. While this increased likelihood of fracture has previously been reported, there is variation in the degree of significance attached to this risk. To date there does not appear to be any previously published data specifically related to femoral fracture and haemochromatosis. The data presented in this paper provides evidence of increased risk, specifically of femoral fracture in this patient group, and given the morbidity and mortality associated with femoral fracture this is a clinically relevant finding.

There are some limitations to consider. UK Biobank participants were healthier than the general population at baseline,^38^ but risk estimates are from incident disease/operations during follow-up so should be less susceptible to this bias at baseline. There is a small chance of misclassification bias for self-reported disease outcomes at baseline, but this is minimised by the fact a trained nurse conducted the interviews with participants. No data were available on ferritin concentrations or transferrin saturation in UK Biobank, so we are unable to examine iron loading within genotype groups. However, there are many sources of error in ferritin levels, and genotyping is the gold standard especially for p.C282Y homozygotes for diagnosing haemochromatosis^39^. With widespread genotyping now becoming available, many patients can now be identified as being at risk of haemochromatosis based on genotype, and this paper has quantified that risk. Also, Kieley et al has previously discussed the non-response of arthritis to de-ironing treatment^20^. We did not exclude those participants with a diagnosis of alcohol dependency, either via self-report or through hospital or primary care data, therefore the risk and incidence of fracture relative to excess alcohol intake needs further research. Also, we did not examine medication use such as corticosteroids or pain medication. Disease diagnosis may be an underestimate as primary care data were only available for a subset of the cohort. In addition, as the mean age of participants was 57 years, and this paper focussed on new incidence of OA the overall burden of haemochromatosis related arthropathy, given its earlier onset, might also be underestimated. Lastly, we studied a European ancestry population so results may not be generalizable to more diverse populations.

## Conclusions

Male p.C282Y homozygotes have an increased risk of OA, hip, knee, shoulder and ankle replacement, osteoporosis and femoral fracture compared to those without mutations. Female p.C282Y homozygotes had an increased risk of OA only. This may have implications for possible earlier diagnosis of haemochromatosis by testing for iron overload and HFE genotypes of at-risk individuals, including at orthopaedic and fracture clinics.

## Ethics approval

The UK Biobank gained ethical approval from the North West Multi-Centre Research Ethics Committee (Research Ethics Committee reference 11/NW/0382). At the baseline assessment, participants gave written informed consent for data collection, genotyping from blood samples, and linkage to electronic medical records for follow-up.

## Author contributions

LRB, KMK, DM and JLA designed the study. LRB and JLA analysed the data and drafted the article. LRB, KMK, LCP, DM and JLA interpreted the data. KMK, DM and LCP revised the article critically for important intellectual content. All authors approved the final version to be published.

## Data availability

Data are available on application to the UK Biobank (https://www.ukbiobank.ac.uk/enable-your-research/register).

## Data Availability

Data are available on application to the UK Biobank

https://www.ukbiobank.ac.uk/enable-your-research/register

## Acknowledgements

This research was conducted using the UK Biobank resource, under application 14631. We thank the UK Biobank participants and coordinators for the dataset. We would also like to thank Professor Patrick Kiely (St Georges University Hospital NHS Trust) for his valuable and constructive suggestions during the writing of this paper. His willingness to give his time has been very much appreciated.

