## Supplementary Information for "Haemochromatosis genetic variants and musculoskeletal outcomes: 11.5 year follow-up in the UK Biobank cohort study"

### Supplementary Methods

#### Genotyping of *HFE* p.C282Y and p.H63D

UK Biobank used Affymetrix microarrays (~800,000 markers directly genotyped). Data were on 451,143 UK Biobank participants of European ancestry with *HFE* p.C282Y (rs1800562) and *HFE* p.H63D (rs1799945) genotype information. *HFE* p.H63D was directly genotyped in the microarray data. *HFE* p.C282Y was not directly genotyped so standard imputation methods were applied.[1] 98.7% of participants were imputed with 100% confidence. 1.26% were recoded (i.e. estimated genotype dose between 0 and 0.25 set to 0, values between 0.75 and 1.25 set to 1, and between 1.75 and 2 to 2) and the remaining 0.04% of participants (n=183) were excluded due to imprecise p.C282Y imputation.

**Supplementary Table 1**. Incident disease/procedure coding from follow-up in hospital inpatient data

| Disease/procedure | ICD-10 Codes | OPCS-4 Codes |
|---|---|---|
| Hereditary haemochromatosis | E83.1* | N/A |
| Osteoarthritis | M15.0; M15.1; M15.2; M15.9; M16.0; M16.1; M17.0; M17.1; M18.0; M18.1; M19.0 | N/A |
| Hip replacement | N/A | W37;W370;W371;W372;W373; W374;W38;W380;W381;W382; W383;W384;W46;W460;W461; W462;W463;W47;W470;W471; W472;W473;W93;W930;W931; W932;W933;W94;W940;W941; W942;W943;O171;O172;O173; W580;W581;W582 |
| Knee replacement | N/A | O18*; W40*; W41*; W42* |
| Ankle replacement | N/A | O32; O320; O321; O322; O323; O324; O325 |
| Shoulder replacement | N/A | O06; O060; O061; O062; O063; O068; O069; O07; O070; O071; O072; O073; O078; O079; O08; O080; O081; O082; O083; O084; O088; O089; O09; O091; O098; O099; O10; O101; O108; O109 |
| Osteoporosis | M80; M81; M811; M812; M813; M814; M815; M816; M818; M819 | N/A |
| Any fracture | S02; S12; S22; S32; S42; S52; S62; S72; S82; S92; T02; T08; T10; T12; T14.2 | |
| Femoral fracture | S72; S72.0; S72.10; S72.11; S72.20; S72.21 | N/A |
| Wrist fracture | S52.5; S52.6 | N/A |

ICD-10 = International Classification of Diseases 10th revision codes; OPCS-4 = OPCS Classification of Interventions and Procedures version 4.

**Supplementary Table 2**. Incident disease coding from follow-up in primary care data

| Disease | READ-2 | READ-3 |
|---|---|---|
| Osteoarthritis | 14G2.; 2G26.; 7P204; N05..; N050.; N0500; N0501; N0502; N0503; N0504; N0505; N0506; N0507; N050z; N051.; N0510; N0511; N0512; N0513; N0514; N0515; N0516; N0517; N0518; N0519; N051A; N051B; N051C; N051D; N051E; N051F; N051G; N051z; N052.; N0520; N0521; N0522; N0523; N0524; N0525; N0526; N0527; N0528; N052z; N053.; N0530; N0531; N0532; N0533; N0534; N0535; N0536; N0537; N0538; N0539; N053z; N054.; N0540; N0541; N0542; N0544; N0545; N0546; N0547; N0548; N0549; N054z; N05z.; N05z0; N05z1; N05z4; N05z5; N05z6; N05z7; N05z8; N05z9; N05zA; N05zB; N05zC; N05zD; N05zE; N05zF; N05zG; N05zH; N05zJ; N05zK; N05zL; N05zM; N05zN; N05zP; N05zQ; N05zR; N05zS; N05zT; N05zU; N05zz; Nyu2.; Nyu20; Nyu21; Nyu22; Nyu24; Nyu25; Nyu27; Nyu28; Nyu29; Nyu2D; Nyu2E | 14G2.; 2G26.; XaLsk; XE1DV; N050.; N0500; XE1DW; XM05w; X76G5; N0502; N0503; X7038; N0505; N0506; N0507; N050z; N051.; N0510; N0511; N0512; N0513; N0514; N0515; N0516; N0517; N0518; N0519; N051A; N051B; N051C; XaEGd; XaEGe; XaEGf; X703A; N051z; N052.; N0520; N0521; N0522; N0523; N0524; X7043; XE1DX; N0526; N0527; N0528; N052z; N053.; N0530; N0531; N0532; N0533; N0534; X7042; XE1DY; N05zJ; N0536; XaYQD; XE1DZ; X703L; N0537; N0538; N0539; N053z; N054.; N0540; N0541; N0542; N0544; N0545; N0546; N0547; N0548; N0549; N054z; XE1Da; N05z0; N05z1; X7034; X7035; XE1Dd; XE1De; XE1Df; N05zL; XE1Dg; X7030; X7031; X702z; N05z8; N05z9; N05zA; N05zB; N05zC; N05zD; N05zE; N05zF; N05zG; N05zH; N05zK; N05zM; N05zN; N05zP; N05zQ; N05zR; N05zS; N05zT; N05zU; Nyu2.; Nyu20; Nyu21; Nyu22; Nyu24; XE1GT; Nyu27; XE1GU; Nyu29; Nyu2D; XE1GV |
| Osteoporosis | 14GB.; 58EG.; 58EM.; 58EV.; 66a..; 9kj..; 9kj0.; 9Od..; 9Od0.; 9Od2.; 9Od3.; 9Od4.; 9Od5.; 9Od6.; 9Od7.; 9Od8.; 9Od9.; N330.; N3300; N3301; N3302; N3303; N3304; N3305; N3306; N3307; N330A; N330B; N330C; N330D; N330z; N3312; N3313; N3314; N3315; N3316; N3318; N3319; N331A; N331B; N331H; N331J; N331K; N331L; N331M; N331M; N331N; N331N; N3746; NyuB0; NyuB1; NyuB2; NyuB8 | XaQgT; XaITW; XaITb; XaPE2; XaISd; XaQR9; XaQRT; XaISP; XaISQ; XaISS; XaIST; XaISU; XaISV; XaISW; XaISX; XaISY; XaISZ; N330.; N3300; N3301; N3302; N3303; N3304; N3305; N3306; N3307; N330A; N330B; XaC12; XaIP4; N330z; N3312; N3313; N3314; N3315; N3316; N3318; N3319; N331A; N331B; XaD2s; XaD4I; XaD4J; XaD4K; XaIIp; XaIIp; XaNSP; XaNSP; N3746; NyuB0; NyuB1; NyuB2; NyuB8 |
